# Age-adjusted associations between comorbidity and outcomes of COVID-19: a review of the evidence

**DOI:** 10.1101/2020.05.06.20093351

**Authors:** Kate E. Mason, Philip McHale, Andy Pennington, Gillian Maudsley, Jennifer Day, Ben Barr

## Abstract

**Background:** Current evidence suggests that older people and people with underlying comorbidities are at increased risk of severe disease and death following hospitalisation with COVID-19. As comorbidity increases with age, it is necessary to understand the age-adjusted relationship between comorbidity and COVID-19 outcomes, in order to enhance planning capabilities and our understanding of COVID-19.

**Methods:** We conducted a rapid, comprehensive review of the literature up to 10 April 2020, to assess the international empirical evidence on the association between comorbidities and severe or critical care outcomes of COVID-19, after accounting for age, among hospitalised patients with COVID-19.

**Results:** After screening 579 studies, we identified seven studies eligible for inclusion and these were synthesised narratively. All were from China. The emerging evidence base mostly indicates that after adjustment for age (and in some cases other potential confounders), obesity, hypertension, diabetes mellitus, chronic obstructive airways disease (COPD), and cancer are all associated with worse outcomes. The largest study, using a large nationwide sample of COVID-19 patients in China, found that those with multiple comorbidities had more than twice the risk of a severe outcome or death compared with patients with no comorbidities, after adjusting for age and smoking (HR=2.59, 95% CI 1.61, 4.17).

**Conclusions:** This review summarises for clinicians, policymakers, and academics the most robust evidence to date on this topic, to inform the management of patients and control measures for tackling the pandemic. Given the intersection of comorbidity with ethnicity and social disadvantage, these findings also have important implications for health inequalities. As the pandemic develops, further research should confirm these trends in other settings outside China and explore mechanisms by which various underlying health conditions increase risk of severe COVID-19.

## BACKGROUND

Since first emerging at the end of 2019, the novel coronavirus SARS-CoV-2 is known to have infected at least 3.7 million people and caused more than 260,000 deaths globally (1). As of the end of April 2020, the pandemic remains uncontained in many parts of the world, and countries recovering from a first wave of infections are concerned about subsequent waves before an effective vaccine is available. To minimise mortality and morbidity in the meantime, and to direct scarce resources most appropriately, a better understanding of the risk factors for progression to severe coronavirus disease 2019 (COVID-19) and death is urgently needed.

Emerging reports and descriptive analyses from China and Italy suggested that people with underlying comorbidities were overrepresented in symptomatic hospitalised cases, and were at increased risk of progression to severe disease and death (2-4). Other countries have reported similar findings as the pandemic has spread (5, 6). Given that the prevalence of comorbidity increases with age, it is unclear whether and how comorbidity independently influences risk of COVID-19 progression. Many early studies into the epidemiology of COVID-19 reported baseline comorbidities of hospitalised patients but not age-adjusted estimates of excess risk associated with comorbidities. Understanding the relationship between comorbidity and COVID-19 outcomes would enhance planning capabilities and potentially our understanding of COVID-19 pathogenesis, management, and prognosis.

To provide timely evidence, we conducted a rapid but comprehensive review addressing the following question: What is the international empirical evidence on the association between comorbidities and severe or critical care outcomes of COVID-19, after accounting for age, among hospitalised patients with COVID-19? Outcomes of interest were:

a. Admission to intensive care unit (ICU)
b. Invasive or non-invasive ventilation
c. Deaths in hospital
d. Progression to severe disease

## METHODS

### Inclusion and exclusion criteria

The inclusion and exclusion criteria focused on comparison of age-adjusted estimates of association between any comorbidity and in-hospital COVID-19 outcomes (severity, critical care or death) (Table 1), in peer-reviewed studies, pre-prints from repositories such as medRxiv, and several grey literature sources of official statistics and evidence summaries, published by 10 April 2020, in English, from any country. We defined comorbidity as a pre-existing health condition present at admission to hospital with COVID-19, including obesity but excluding health-related behaviours such as smoking.

**Table 1.** Review inclusion and exclusion criteria: What is the association between comorbidities and age-adjusted severe or critical care outcomes in hospital patients with COVID-19?

|  | INCLUDE | EXCLUDE |
| --- | --- | --- |
| **Population** | Adult COVID-19 hospital patients.<br>Studies with 10 or more patients. | Not population-based sample (e.g. samples in clinical trials, samples from cruise ships, familial clusters; studies with less than 10 patients).<br>Non-COVID-19 patients (e.g. other pneumonia cases).<br>Community cases not receiving care in hospitals, including general population estimates of the spread of COVID-19.<br>Studies focusing solely on infants and children (not part of a study including adults). |
| **Interventions/outcomes** | Relative risk, hazard ratio, odds ratio associated with comorbidity (pre-existing condition) status on admission, of:<br>i. admission to intensive care<br>ii. invasive or non-invasive ventilation<br>iii. progression to severe disease<br>iv. death in hospital<br>v. composite indicators of i-iv,<br>for any reported comorbidity (chronic illness, pre-existing condition). | Other treatments inside and outside critical care departments, e.g. rates of patients receiving oxygen supplementation. |
| **Comparison** | Patients with and without any comorbidity at admission to hospital. Comorbidity was defined as pre-existing health conditions present at admission to hospital with COVID-19, including obesity. | Comparisons within a sample of patients who all have a comorbidity (e.g. studies of cancer patients only). Comparisons between groups of people based on their health-related behaviours (e.g. smoking), ethnicity or socioeconomic circumstances. |
| **Study design** | All primary quantitative empirical observational studies reporting multivariable regression models (with adjustment for age) or age-specific association between comorbidity and outcome. | Qualitative studies.<br>Intervention studies (e.g. clinical trials of new treatments for COVID-19).<br>Projections or estimations of potential outcomes.<br>Non-empirical studies, including editorials, opinions, or discussion pieces.<br>Studies that do not report comorbidity-related risk estimates<br>Review-level evidence |
| **Publication characteristics** |  |  |
|  | INCLUDE | EXCLUDE |
| **Publication stage, type** | Pre-prints, peer-reviewed publications, grey literature on empirical evidence (e.g. official statistics). | Not applicable. |
| **Language** | English language publications. | Non-English language publications (not available for full text). |
| **Date** | Studies with data from December 2019 and published by 10th April, 2020. | Studies before December 2019. |

**Table 2.** Extracted age-adjusted estimates (and 95% confidence intervals) of excess risk of progression to severe disease or death associated with comorbidities among hospitalised COVID-19 patients (published by 10 April 2020, all from China)

| Comorbidity<br>(ref=comorbidity not present) | Composite endpoint:<br>ICU admission, invasive<br>ventilation, or death | Severe disease (including ICU<br>admission) | Death | Samples sizes |
| --- | --- | --- | --- | --- |
| *Any comorbidity* |  |  |  |  |
| One or more |  | OR=1.83 (0.50, 6.75) <sub>s, B</sub> (11) o |  | 249 |
| One only | HR=1.79 (1.16, 2.77) <sub>sm</sub> (12) N |  |  | 1,590 |
| Two or more | HR=2.59 (1.61, 4.17) <sub>sm</sub> (12) N |  |  | 1,590 |
| *Obesity* |  |  |  |  |
| Overweight |  | OR=1.78 (1.00, 3.21) (10) o |  | 387 |
| Obesity |  | OR=3.35 (1.47, 7.63) (10) o |  | 387 |
| *Cardiovascular and related* |  |  |  |  |
| Hypertension | HR=1.58 (1.07, 2.32) <sub>sm</sub> (12) N | OR=2.71 (1.32, 5.59) <sub>s,t</sub> (14) o |  | 1,590, 487 respectively |
| Coronary heart disease |  |  | OR=2.14 (0.26, 17.79) <sub>B</sub> (16) H | 171 |
| Cardiovascular disease |  |  | HR=1.40 (0.65, 3.03) <sub>c, B</sub> (13) H | 416 |
| Cerebrovascular disease |  |  | HR=1.71 (0.71, 4.09) <sub>c, B</sub> (13) H | 416 |
| Diabetes mellitus | HR=1.59 (1.03, 2.45) <sub>sm</sub> (12) N |  | HR=2.84 (1.01, 8.01) <sub>c</sub> (15) H<br>HR=0.75 (0.38, 1.50) <sub>c, B</sub> (13) H | 1,590, 258<br>416 |
| Cancer | HR=3.50 (1.60, 7.64) <sub>sm</sub> (12) N |  | HR=0.82 (0.18, 3.65) <sub>c, B</sub> (13) H | 1,590, 416 |
| COPD | HR=2.68 (1.42, 5.05) <sub>sm</sub> (12) N |  | HR=0.39 (0.04, 3.68) <sub>c, B</sub> (13) H | 1,590, 416 |

ICU = Intensive care unit
COPD = Chronic obstructive pulmonary disease
HR=hazard ratio
OR=odds ratio
All estimates adjusted for age. Additionally adjusted for:
sm = smoking
s = sex
t = time from symptom onset to admission
c = other comorbidities
B = clinical biomarkers (e.g. of inflammatory response or organ function) – potential intermediate markers of COVID-19 severity and not a reliable estimate of the comorbidity-associated relative risk of the outcome.
Study population in China: N=nationwide, non-Hubei; H=Hubei province; O=other province

### Search strategy and study identification

The search strategy had four arms (Figure 1) involving electronic database searches and a wide range of ‘supplementary’ systematic review search methods.

**Figure 1.**
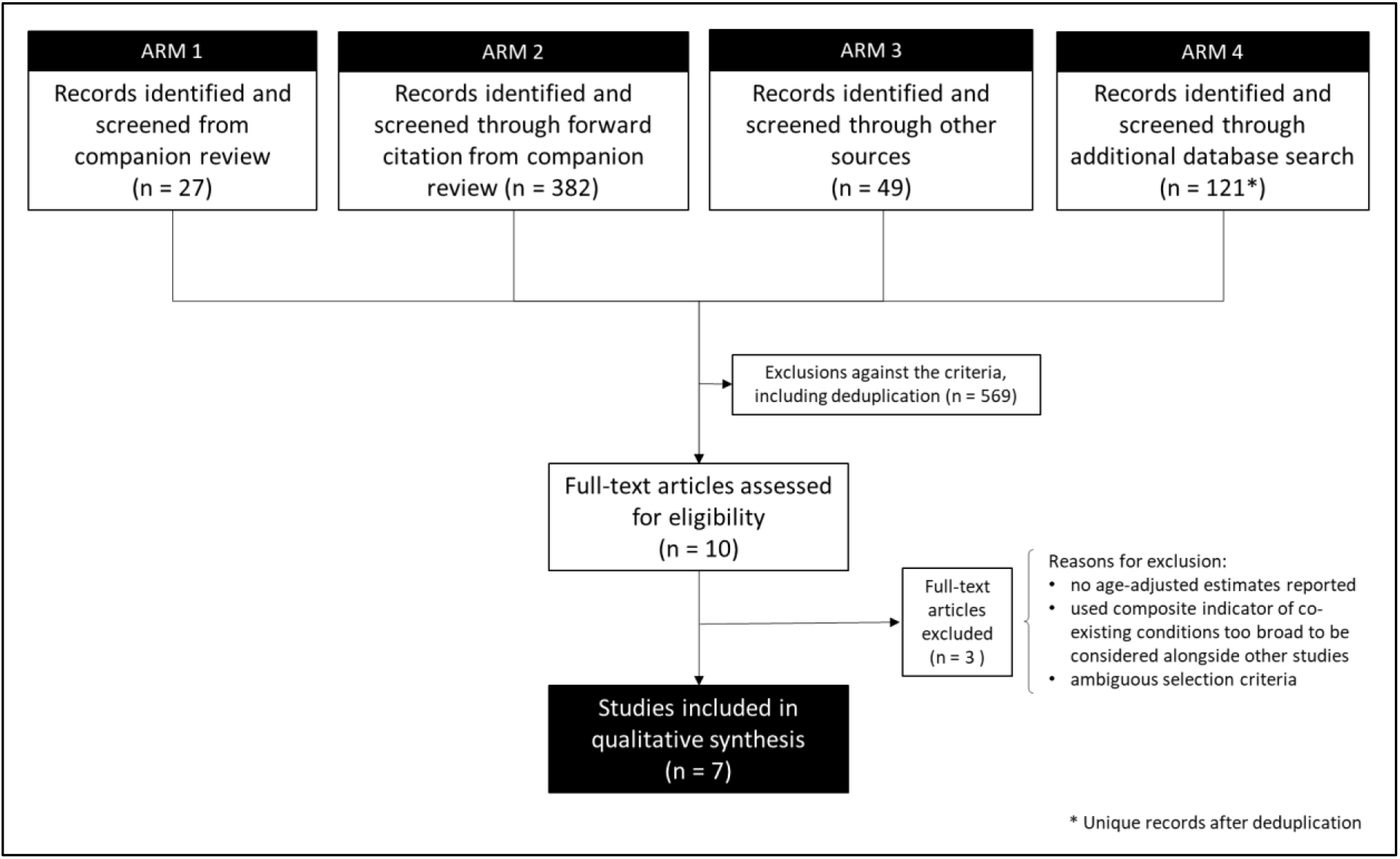
Flow chart of progression of studies through the review of age-adjusted associations between comorbidity and outcomes of COVID-19.

First, we screened the studies already identified in the initial search from our companion review on COVID-19 critical care outcomes (Pennington et al. unpublished, which screened 2665 items), for any that met our narrower search criteria.

Second, all studies identified in that companion review underwent Web of Science and Google Scholar forward-citation searches, with initial filtering for key terms relating to comorbidity and age.

Third, we searched a range of additional sources including the World Health Organization (WHO); communicable disease centres of the USA, Europe, and China; and several COVID-19-specific evidence resources online (shown as “outside sources” in the flow chart, Figure 1; see Appendix 1 for details).

Fourth, in a new search of the MEDLINE *full-text* database (as title and abstract often omit age-adjustment), we modified the companion review’s search strategy to identify additional analytical (rather than descriptive) studies that were focused on comorbidities specifically or reported multivariable analysis of risk factors for severe or critical care outcomes of COVID-19 (see Appendix 1 for full search terms).

### Screening

In each of Arms 1-3, title-abstract screening was followed by full-text screening by a single reviewer. In Arm 4, title-abstract screening by one reviewer excluded any studies clearly not meeting the inclusion criteria, followed by independent title-abstract screening of the remaining studies by two reviewers in EPPI Reviewer-4 systematic review management software (7).

Duplicates remaining after the screening of the additional MEDLINE search were excluded at this stage. To produce this report quickly, three reviewers shared searching and screening tasks, rather than repeating tasks independently, except where otherwise stated. Two reviewers independently screened the full text of the final set of studies screened as potentially eligible.

### Data extraction

One reviewer extracted any age-adjusted estimate of excess risk (relative risk, hazard or odds ratio) associated with any comorbidity (variously defined) for the outcomes of interest, recording them in an Airtable Pro database. A second reviewer checked each extraction for accuracy and missing data. Where studies reported multiple estimates adjusted for different sets of covariates (e.g. age alone, age plus sex, other comorbidities, smoking status), all relevant age-estimates were extracted. One reviewer then selected the most appropriate one for reporting in the review, checked by others. No assessment was made at this stage of the comparability of estimates or the appropriateness of the covariate selection in each study’s adjusted regression models. Instead, such assessment was made in the synthesis of results.

### Synthesis

Evidence was synthesised narratively (8, 9) and study quality was assessed informally. Limited replication of studies for any individual comorbidity and the inclusion of non-peer-reviewed preprints meant that meta-analysis was not viable.

## RESULTS

A total of 579 studies were screened (Figure 1), of which seven studies were included in the review (10-16) (Appendix 2).

No studies were identified in the screening of 27 studies identified from the companion review. Forward citation searches of those 27 led to the screening of 382 studies and identified three primary studies for potential inclusion though (10, 15, 17). One of these was then excluded because it was not clear if selection into the study was limited to hospitalised cases (17). Of 49 studies screened from outside sources, several reviews were identified that led to the identification of five primary studies of potential relevance, of which four were assessed as eligible for inclusion (11, 13, 14, 16). A Centers for Disease Control and Prevention (CDC) publication (18) was excluded because it analysed a composite indicator of ‘underlying health conditions’ that also included health-related behaviours. The additional MEDLINE search identified 121 unique studies and excluded 81 after screening of titles and abstracts. From the remaining 40 studies, two not already identified through other avenues were identified as potentially eligible for inclusion (12, 19). One of these was excluded because the age-adjusted estimates anticipated by the methods were not reported (19).

Of the seven papers, five had been peer-reviewed (11-14, 16) while two were pre-prints not yet peer-reviewed (10, 15).

### Characteristics of the included studies

All the included studies were based in China during the initial stages of the pandemic. Three used data from Wuhan and the Hubei province (13, 15, 16), the epicentre of the Chinese outbreak, while the others focused on cases hospitalised outside Hubei, using either national samples (12) or records from a single tertiary hospital in another province (10, 11, 14). Sample sizes ranged from 171 (16) to 1,590 (12).

Our review identified mostly studies with a cohort design, although only four studies accurately specified the study design. Convenience sampling seemed to be most common, however two studies only stated recruitment of consecutive patients (13, 15) and only two described excluded cases (13, 16). Sample construction in other studies was not explicit. Only one study specified inclusions and exclusions (13), albeit without comparing the 41% inclusions with the excluded cases.

One study used a composite endpoint (death or admission to ICU or invasive ventilation) as the outcome (12). Three studies recorded death as the primary outcome (13, 15, 16), and three severe disease, including ICU admission (10, 11, 14). In one study, severity was not clearly defined but appeared to be based on a clinical evaluation of CT scans (14). In another study (15), severe and critical COVID-19 illness was clearly defined based on the diagnostic and treatment guidelines of the National Health Committee of China.

Comorbidities analysed were obesity, diabetes mellitus, hypertension and other circulatory disease (including cardiovascular and cerebrovascular disease), cancer, and chronic obstructive pulmonary disease (COPD). Additionally, two studies (11, 12) reported the association of one of the outcomes with the presence of any (or multiple) comorbid conditions rather than, or as well as, specific conditions. In five studies, information on comorbidities was collated from medical records but two of these did not specify the exact method. One study (13) reported details of a robust method of two researchers collecting data from medical records, followed by independent review and data entry by two analysts. Another study (16) reported using a structured form adapted from the WHO/International Severe Acute Respiratory and Emerging Infection Consortium case-record form to record clinical data, which two physicians then checked. One study included comorbidities self-reported on admission (12), and another measured weight and height on admission to calculate body mass index (BMI) (10).

All included studies adjusted for age using multivariable regression models (Cox (n=3); logistic (n=3) or unspecified (n=1)). Most studies also adjusted for additional covariates, but the particular extra covariates differed across studies. In some studies (13, 14), details of the adjustment set were ambiguous, undermining confidence in any interpretation of the estimate. Additional covariates included sex, smoking, other comorbidities, and time from symptom onset to hospital admission. Importantly, four studies (11, 13, 15, 16) reported estimates of excess risk associated with comorbidity adjusted for clinically ascertained biomarkers (such as inflammatory response or organ function). In these four studies, full results are reported from multivariable models, and it appears (though is not stated explicitly) that comorbidities are considered potential confounders of associations between clinical predictors and disease progression. This is in contrast with the other three studies (10, 12, 14), which focused on estimation of the independent risk of experiencing a severe outcome *according* to comorbidity status, adjusted for age and other confounders.

In the remainder of the narrative synthesis, we report first and in most detail on larger studies and those studies that directly attempted to estimate the age-adjusted effect of a comorbidity on any of the outcomes. Studies presenting models not appropriately designed for answering that particular research question are noted but considered less relevant.

### Comorbidity and severe/critical care COVID-19 outcomes

#### Any comorbidity

From the small number of studies published to date, there is preliminary evidence to indicate that hospitalised patients with any comorbidity are more likely to be admitted to ICU, require invasive ventilation, or die from COVID-19. In a nationwide study of 1,590 patients, Guan et al (12) examined the effect of ‘any comorbidity’ – as well as specific ones – and found evidence of a dose-response relationship. After adjustment for age and smoking status, patients with a single comorbidity had a 79% greater hazard of experiencing a severe outcome than patients with no comorbidities, while those with multiple comorbidities had a 159% increased hazard of severe outcome (HR=2.59, 95% CI: 1.61-4.17) (12). A second, smaller study of 249 patients in a Shanghai hospital estimated an age-adjusted odds ratio of similar magnitude for any comorbidity, but its wide 95% confidence interval included the null (0.50-6.75) (11). That study adjusted for clinical biomarkers, potentially making this an underestimate of the direct effect of comorbidity on the outcome.

#### Overweight and Obesity

Cai et al (10) examined the relationship between overweight and obesity and progression to severe pneumonia in 387 hospitalised COVID-19 cases in Schenzen, a Chinese city outside the Hubei province. They found that, independent of age, obesity significantly increased the risk of developing severe pneumonia in COVID-19 patients, compared with patients of normal weight (OR=3.35, 95% CI: 1.47-7.63). A dose-response relationship was observed, with overweight patients at intermediate risk (OR=1.78, 95% CI: 1.00-3.21). They also observed that the relationship was particularly pronounced in men (HR=5.40, 95% CI 1.93-15.09).

#### Hypertension and CVD

In their large nationwide study, Guan et al also examined a range of specific comorbidities and found that, after adjusting for age and smoking status, patients with hypertension at admission were 58% more likely to reach the composite endpoint (ICU admission, invasive ventilation, or death) than those without hypertension (HR 1.58, 95% CI: 1.07-2.32) (12).

Similarly, Shi et al (14) found that the presence of hypertension at hospital admission was associated with 2.71 times the odds of severe disease (ICU admission), in a retrospective cohort of 487 patients in Zhejiang province of China, after adjusting for age, sex, and time from symptom onset to admission. Ambiguous reporting of the adjustment set for this analysis meant that this estimate should be interpreted with some caution.

The only studies reporting hazard ratios for other cardiovascular and related comorbidities used multivariable models adjusted for clinical biomarkers that could relate to mediators of the pathway from prior comorbidity to COVID-19 severity (13, 16). Estimates are thus unlikely to represent the independent risk of severe outcomes associated with having these comorbidities. Their direction is consistent with an increased relative risk of death, but the confidence intervals are wide.

#### Diabetes mellitus

In studies examining diabetes mellitus as a comorbidity, the authors did not distinguish between Type 1 and Type 2 diabetes. The large, nationwide study (12) found that hospitalised COVID-19 patients with diabetes had a 59% increased risk of the composite endpoint (ICU admission, invasive ventilation, or death) (HR 1.59, 95% CI: 1.03-2.45), after adjusting for age and smoking status. Zhang et al (15) in their study of 258 COVID-19 patients at a Wuhan hospital also found that those with diabetes were more likely to die in hospital, and point estimates were not materially different when adjusted for additional comorbidities as well as age (HR=2.84 cf. 2.80). After additionally adjusting for clinical biomarkers and other comorbidities, Shi and colleagues (13) reported no significant association between diabetes and mortality (HR=0.75; 95% CI: 0.38-1.50).

#### Cancer

Guan et al (12) also examined cancer as a comorbidity, finding a substantially elevated risk of their composite endpoint amongst patients with cancer at admission to hospital. After adjusting for age and smoking status, patients with cancer had 3.5-fold the hazard of ICU admission, invasive ventilation, or death in hospital compared with patients without cancer (95% CI: 1.60-7.64). Just as for diabetes, Shi et al (13) reported no significant association between cancer and mortality, after additionally adjusting for clinical biomarkers and other comorbidities (HR=0.82; 95% CI: 0.18-3.65).

#### COPD

Regarding COPD, Guan et al found that hospitalised patients with COPD had 168% higher risk of reaching that study’s composite endpoint (ICU admission, invasive ventilation, or death) than patients without COPD (HR=2.68, 95% CI: 1.42-5.05), adjusted for age and smoking status. Again, Shi et al (13) reported no significant association between COPD and mortality after additionally adjusting for clinical biomarkers and other comorbidities (HR=0.39; 95% CI: 0.04-3.68).

## DISCUSSION

### Summary of the findings

There is limited research on comorbidities as independent risk factors for severe COVID-19, but the emerging evidence base we identified supports the hypothesis that various underlying health conditions confer additional risk of severe disease and mortality among people hospitalised with COVID-19. Obesity, diabetes mellitus, hypertension, cancer, and COPD were all significantly associated with severe outcomes in the studies well designed to assess those associations – at least 50% higher than for people without the comorbidity. A dose-response relationship was reported for multiple comorbidities and for overweight and obesity.

Comorbidity has previously been shown to be associated with elevated risk of worse clinical outcomes in other severe acute respiratory outbreaks such as SARS (severe acute respiratory syndrome), MERS (Middle East respiratory syndrome), and avian influenza (20-22). The findings of this review are consistent with the hypothesis that comorbidity also predisposes individuals to poorer outcomes in this current COVID-19 pandemic. Whilst the mechanisms remain poorly understood, there are numerous biologically plausible explanations. The pathogenesis of severe COVID-19 is thought to involve dysregulated proinflammatory immune response and subsequent multi-system damage (23). Many underlying conditions may leave affected individuals more vulnerable to the effects of this. Obesity, for example, tends to reduce lung function and dysregulate the immune system (24). Similarly, diabetes mellitus can impair immune function (25), as do many cancer treatments. Patients with pre-existing hypertension and other CVD may be at heightened risk of severe outcomes through various mechanisms, including therapeutic upregulation of ACE2 (the host receptor for SARS-CoV-2), and greater vulnerability to hyperinflammatory immune responses and cardiac complications that are common with severe COVID-19 (26, 27).

### Strengths and limitations in the evidence base

The review includes two studies yet to undergo peer review, so these must be treated with caution, but it was deemed important to include these given the emerging pandemic and the need for timely evidence reviews.

Our review identified mostly studies with a cohort design, appropriate for identifying independent risk factors for an outcome, although only four studies accurately specified the study design. Details of sampling and inclusion and exclusion criteria were scant in some studies, which undermines generalisability, and there may be a risk of selection bias when making inferences about the hospitalised COVID-19 population. Furthermore our review is limited to the hospitalised population of COVID-19 cases. Our conclusions therefore only indicate the increased risk associated with comorbidities in hospitalised patients; we do not know what effect comorbidities have on the initial risk of being admitted. Compared with many earlier case-series reports from China (Pennington et al, unpublished), the studies we identified had relatively large sample sizes, providing adequate statistical power to detect differences between groups of patients with and without comorbidities.

Outcomes and definitions varied markedly between studies, compromising comparison of results and pooling estimates. Furthermore, understandably, patients in many studies had not yet reached their clinical endpoint and so the results are not complete. Many papers did not clearly specify the methods for data collection, particularly for recording comorbidities. In some, lack of rigour in comorbidity ascertainment might have led to misclassification and possible bias. Where case definition was not specified, comparing different studies was difficult. For example, some studies included cardiovascular disease or cerebrovascular disease as a comorbidity without specifying conditions included or excluded or specifying diagnostic criteria. Similarly, no study was clear about whether it distinguished between Type 1 and Type 2 diabetes mellitus, despite the two disease types having distinct aetiologies, age profiles (juvenile onset vs older ages), manifestations, and treatments, and therefore potentially not conferring equivalent risk with respect to COVID-19 severity.

To analyse the association between the comorbidity and the outcome, all studies used multivariable regression analyses. Across studies, otherwise similar models differed considerably in adjusting for obvious confounders, such as sex or smoking, making comparison challenging. Furthermore, our inclusion and exclusion criteria retained studies reporting multivariable models that contained comorbidities as potential confounders of associations between clinical predictors and disease progression. In those studies, it is possible that the clinical predictors are intermediates on pathways through which comorbidity elevates the risk of death from COVID-19. If so, interpreting the hazard/odds ratios for comorbidity from these models would lead to a bias towards the null in any estimated effect of comorbidity, due to overadjustment by potential mediators.

Overall the seven studies varied considerably both in the quality of the design and reporting. Whilst hasty research and publication are understandable in a global pandemic, rigour should not be compromised, as London & Kimmelman recently argued (28). Indeed, there is an ethical imperative to ensure that the conduct and reporting of research in a pandemic crisis maintains high standards of validity, reliability, and integrity to provide sufficiently robust evidence to inform clinical practice and public health policy.

### Strengths and limitations of the review

This review has been rapid and timely, while also being as comprehensive as possible within those limits. A companion review provided forward citations, and our full-text MEDLINE search included pre-print archives as well as peer-reviewed literature, reflecting the fast-moving early stages of the pandemic and the increasing use of pre-print archives. While each early step used a single reviewer, full-text screening used double screening. While we did not conduct a formal quality assessment, we have outlined here the major quality issues to consider in interpreting the results.

The timing of the review and inevitable delay between research and publication meant that only studies from China met the criteria for inclusion. The Chinese study patients may be healthier because of different criteria for admission compared with other countries (e.g. the United Kingdom), where only relatively serious cases are hospitalised. The comorbidity profile of China also differs from that of many other parts of the world (29-31). Future studies must therefore examine comorbidities as risk factors for progression to severe COVID-19 and death in other settings globally. Updating this review to include such subsequent studies from outside China would be a valuable next step. Since the search completed in mid-April, a large study of hospitalised cases in the UK has emerged in pre-print (32). While it was thus not included in our analysis, it reports age-and-sex-adjusted estimates of mortality associated with a range of comorbidities that are consistent with those from the Chinese studies. They found higher inhospital mortality associated with obesity, cancer, chronic cardiac, pulmonary and kidney disease, and also dementia.

### Implications for future data collection and research

COVID-19 studies to date have been produced very rapidly in a fast-moving pandemic. Reporting of methods would seem to have been adversely affected, and these preliminary studies are likely to feed into reviews and inform policy decisions. Transparent and detailed reporting of methods is required for accurate interpretation, particularly a clear rationale for model adjustment. Pre-agreed consistent methods of sample selection, description, and design (including variables to be measured and data collection tools) would facilitate more effective use and application of research efforts. It would also enable pooling of results from different locations and settings to provide high-quality evidence as quickly as possible.

### Implications for policy and practice

People with various comorbidities appear, from the emerging evidence, to be at increased risk of severe disease progression and death after developing COVID-19. Elevated risk does not appear to be limited to specific comorbidities or organ systems. Rather, many of the most common chronic conditions confer an elevated risk of severe outcome, and there is also evidence that multimorbidity adds further risk. Given the relatively high prevalence of most of the comorbidities covered in this review, the implications of elevated risk among those affected are substantial. A recent study estimated that one in five individuals globally may be at increased risk of severe COVID-19 due to underlying conditions (33). This is likely to be an underestimate, as the study did not include obesity.

Whether COVID-19 accelerates the underlying condition, or weakened underlying organs or immune response increase vulnerability to severe COVID-19, or both, remains an important area for further research. Understanding the mechanisms involved is critical. Nevertheless, even without a full explication of the mechanisms, epidemiological evidence of an association between comorbidities and poor in-hospital outcomes still supports action to protect these groups and mitigate their elevated risk.

In terms of primary prevention of COVID-19, the increased risk associated with many comorbidities supports strong, targeted measures to ‘shield’ people with comorbidities, and it suggests a need for public health campaigns to promote awareness of these elevated risks to enable people to protect themselves appropriately. If a vaccine becomes available, prioritisation of vaccination should be based on risk. In terms of secondary prevention of COVID-19, early COVID-19 detection must be promoted amongst those with comorbidities, to take advantage of any effective treatments that are developed. In terms of tertiary prevention, evidence of elevated risk of severe outcomes can also inform decisions around triage, patient management, treatment (prioritisation and care provision). It may also point to a need for differential approaches to the care of recovered and recovering COVID-19 patients with comorbidities, due to their additional needs. In terms of resource allocation, the evidence of increased risk associated with comorbidities has implications for healthcare system demand in areas of high comorbidity prevalence. To address the greater burden of COVID-19 in communities with more preexisting conditions, greater resources will be needed in these communities. Current approaches, however, are not sufficiently taking into account the higher levels of need experienced by some communities due to comorbidities (34). This emerging evidence also has implications for the preparation for second and subsequent waves of community transmission. In addition to careful management of people with chronic conditions to minimise risk over the longer reach of this pandemic, it also highlights an added urgency for reducing the prevalence and incidence of comorbidities, through greater support for prevention efforts and addressing the wider determinants of health.

Finally, the intersection of underlying comorbidity with socioeconomic disadvantage, geography, and demographic factors, especially ethnicity, may prove to be a potent mix that will lead to a widening of health inequalities. Recent data from the Office of National Statistics in the UK also showed that COVID-19 mortality rates are more than twice as high in the most deprived parts of England than in the least deprived areas (35). Deprivation increases the risk of poor health; higher levels of comorbidity in more disadvantaged groups is likely to be driving some of these COVID-19 inequalities, but social factors may also be playing a role (such as overcrowded housing, employment in essential occupations, particularly public-facing roles or others where physical distancing is not feasible, and greater reliance on public transport). Also in the UK, people from ethnic minority backgrounds are overrepresented among deaths from COVID-19 (36), with evidence that ethnicity is a risk factor independent of deprivation (37). Some of this disparity is likely to be due to the higher prevalence of common comorbidities in some of these groups. People with chronic health conditions, already disadvantaged and underrepresented in the workforce, are also more likely to be disadvantaged by the control measures. Without an effective vaccine or treatment many are likely to be isolated or shielded, and therefore unable to conduct normal activities of daily living and working, for the foreseeable future. This has potential lasting implications for their financial, social, and mental wellbeing. Our review highlights that there are potentially people with chronic conditions across all age groups, such as younger people with Type 1 diabetes, conflated with older people. As far as is possible, people need support tailored to their need. Rather than being a ’great leveller’, this pandemic highlights the potential consequences of existing health inequalities and uneven distribution of underlying health conditions. Without concerted effort, the COVID-19 pandemic may lead to widening health inequalities between social, ethnic, and geographical groups. Responses to the pandemic must therefore prioritise and mitigate the unfair burden shouldered by disadvantaged and ethnic minority groups.

### Conclusion

Building on evidence that people with comorbidities were overrepresented in hospitalised cases of COVID-19, this review compiles estimates from age-adjusted regression modelling across seven studies from China. It summarises for clinicians, policymakers, and academics the most robust evidence to date on this topic, to inform patient management and resource allocation for tackling the pandemic. Given the intersection of comorbidity with ethnicity and social disadvantage, these findings also have important implications for health inequalities. As the pandemic develops, further research is required to confirm these trends in other settings outside China and to explore the mechanisms by which various underlying health conditions increase risk of severe COVID-19.

## Data Availability

Review article. All search details included in supplementary material.

## Author contributions

BB, AP, KM, and PM conceived of and designed the review. KM, PM and AP undertook the searching and screening, with search support from the companion critical care review team. KM did the initial data extraction and summarised study characteristics. GM checked data extraction for accuracy and missing data. JD completed an informal quality assessment of the included studies. All authors contributed to the writing of the manuscript.

## Ethics committee approval

None required

